# General psychopathology and substance use disorders: An indicator of functional impairment

**DOI:** 10.1101/2025.11.24.25340502

**Authors:** Asha Pavuluri, Glenda Mello, Devin Butler, Kristi Carrasquillo, Cristina M. Risco, Carl W. Lejuez, Norman B. Schmidt, Brian M. Hicks, Kristian E. Markon, Robert F. Krueger, Edward M. Bernat

## Abstract

**Introduction:** Research represented in the Hierarchical Taxonomy of Psychopathology (HiTOP) suggests that broader, dimensional constructs of Internalizing (**INT**; Fear and Distress), Externalizing (**EXT**; Disinhibition and Antagonism), and Thought Disorder (**TD**) measures better characterize the structure of common mental health disorders. Shared variance across these dimensions has long been noted, suggesting an even broader general psychopathology factor (***p*-factor**). Polysubstance use (**PSU**) is defined as the use of two or more addictive drugs simultaneously or concurrently and polysubstance use disorder (**PSUD**) is the presence of two or more substance use disorders (**SUDs**). PSU is associated with broad increases across problem behaviors and psychopathology. **Objective.** The goal of the present project was to identify the relationship between different underlying HiTOP factors and SUDs/PSUDs across a treatment sample and a nationally representative sample.

**Methods:** The first sample contained 2617 participants from a residential substance use treatment center in the DC area, serving primarily low-income, African American population. The second sample, National Comorbidity Survey-Replication (NCS-R), included 5,692 adults, aged 18 and older, selected using a multistage stratified cluster sampling design representative of the US household population (Kessler et al. 2004). To accomplish the primary objective, SUDs were separated from the rest of the DSM diagnoses, forming a separate SUDs/PSUDs latent factor. Then the HiTOP model was constructed from the remaining diagnoses leaving the subfactors TD, Fear, Distress, and Antagonism, in addition to the *p*-factor. Structural equation modeling, completed in MPlus software, was used to develop the latent HiTOP and SUDs/PSUDs factors and conduct mediation models.

**Results:** Direct effects indicated robust relationships between SUDs/PSUDs and the *p*-factor, as well as each of the sub-factors. Mediation models indicated that *p*-factor fully mediated the relationship between SUDs/PSUDs and TD and Distress, as well as the majority of the relationship between SUDs/PSUDs and Fear and Antagonism. Indirect effects indicated that the relationship between Antagonism and SUDs/PSUDs remained significantly positive relative to *p*-factor as a mediator, and Fear maintained some negative prediction in the treatment sample but was fully mediated in the NCS-R data.

**Discussion:** Concisely, there is a similar and robust relationship between the latent HiTOP *p*-factor measure and SUDs/PSUDs across the samples. Beyond the broad *p*-factor direct effects accounting for most of SUDs/PSUDs, indirect effects indicate that there is some unique contributions to SUDs/PSUDs from Antagonism (positive) and Fear (negative). Results support the inference of a practically important and statistically robust relationship between mental health and substance use, as well as the presence of smaller differential contributions to SUDs/PSUDs from INT and EXT.

## Introduction

The Hierarchical Taxonomy of Psychopathology (HiTOP)^1^ model is a widely studied transdiagnostic system that helps provide a model of comorbidities among problem behaviors. We aimed to characterize substance use disorder (SUD) comorbidities in the context of the HiTOP model.

Comorbidity between psychopathology and substance use (SU) has been observed and investigated for many years^2–4^. Elevated rates of SU have been observed across person with depression, bipolar, anxiety, schizophrenia, and antisocial and borderline personality disorders)^5–7^. However, due to strong associations between SUDs and Externalizing psychopathology, comorbidities between SUDs and the other dimensions in HiTOP (i.e., Thought Disorder and Internalizing, which includes depression, anxiety, and PTSD) have been understudied. The approach we took was to separate SUDs out from HiTOP, and then characterize the relationship between SUDs and the transdiagnostic dimensions in HiTOP.

### HiTOP Transdiagnostic Framework and Substance Use

The HiTOP framework models comorbidity by accounting for the shared variance across psychiatric diagnoses via latent factors using structural modeling^1^. Within this model are a series of spectra (sometimes referred to as super factors), including Internalizing (INT; e.g., Distress disorders: depression, anxiety, PTSD or Fear disorders: panic, obsessive compulsive disorder), Externalizing (EXT; e.g., SUDs: alcohol use disorder, cocaine use disorder or antagonistic disorders: borderline personality disorder, ADHD), Thought Disorders (TD; e.g., schizophrenia, psychosis), and an overarching general psychopathology factor (*p*-factor) representing the shared variance across INT, EXT, and TD (among other factors, which are not represented in detail in the present project). HiTOP latent factors predict that related disorders within a dimension are more likely to occur concurrently and sequentially in time different than it is for individuals with no problems in that dimension^8^. For example, a person with anxiety as an adolescent may be more vulnerable to depression as a young adult, and posttraumatic stress if exposed to trauma^9^.

SUDs have been assessed in this framework and the strong comorbidities of substance use with disinhibitory psychopathology (e.g. antisocial behavior) has driven HiTOP models to include SUDs as part of the EXT dimension. Substantial research supports this, where evidence pointing to a broad undercontrolled-disinhibitory (cf. behavioral disinhibition) developmental pathway to SUDs is strong and has been well-documented^10,11^.

Notably, a behavioral-inhibition developmental pathway for internalizing behaviors has also been identified, analogous to the behavioral-disinhibition pathway to externalizing behaviors noted above, with emerging evidence that it too is associated with increased substance use^12^. Broad evidence supports this, where individual internalizing diagnoses and behaviors have been widely associated with substance use, with some work proposing a separable internalizing pathway to SUDs^13^. First, comorbidity between alcohol use and depression is well-documented^14–16^. PTSD has also been well-assessed as related to increased SU, both for alcohol and other substances^17^. In some cases, PTSD precedes and serves a causal role in SU^18^, and those with comorbid PTSD and SU are associated with worse outcomes^19^. Anxiety has similarly been well-related to SU^20^, with both sedative and psychostimulant substances being used^21^. Alcohol use for those with social anxiety has been particularly noted, as a self-medication^22^, but the causes related to different anxiety disorders and different substances of abuse are still in debate^23^.

Finally, TD has also been reliably related to SU^24–27^, including alcohol, nicotine, and other illicit substances, where TD symptoms can relate to increased substance use and substance use can trigger TD. Given the broad associations between psychopathology and substance use, we aimed to create a comprehensive model the relationships between SUDs and HiTOP dimensions.

Thus, while SUDs has been notably associated with EXT in the HiTOP, it is clear that there are strong and additional associations with both INT and TD. By understanding these relationships, a more detailed model of SU relationships with psychopathology can be developed, and potentially treatment methods, as well. The present study aimed to characterize associations between SUDs and multiple dimensions in the HiTOP framework.

### Substance Use, Polysubstance Use, and Use Severity

Drug overdose deaths generally involve multiple substances^28^, and a majority of those seeking treatment for an SUD have 2 or more SUDs, with some reports suggesting as much as 3.5 average substances for those in treatment for SUDs^29–31^. Polysubstance use disorder (PSUD) is defined as the presence of more than one SUD (simultaneously or sequentially) and is associated with worse course and outcomes of SUDs^32–34^. Consistent with this, a study of 472,624 veterans indicated that increasing PSUDs (1, 2-3, to 3+) were more likely to identify as Black, be homeless, and have hepatic disease^33^. Those with PSUD also had higher rates of schizophrenia, bipolar, major depressive, and personality disorders, as well as greater use of psychiatric inpatient, residential, and rehabilitative treatment and multiple psychotropic medication fills^33^. In a 3-year longitudinal study, comparing polysubstance users who used more substances had poorer physical health and mental health and increased medical and treatment utilization^35^. Tracking across 24 states and Washington, D.C., nearly half (48.9%) of 16,236 overdoses from January through June 2019 involved two or more substances^36^. Broadly, having one SUD increases likelihood for PSUD, and PSUD is associated with worse outcomes, greater severity and functional impairments. Together, current evidence suggests a gap in the current literature in characterizing the relationship between PSUDs and mental health diagnoses as characterized in the HiTOP, which the present project seeks to address.

### Health Disparities

The literature on health disparities indicates differential patterns and prevalence rates of both psychopathology and substance use among diverse and underrepresented populations including racial and ethnic minorities^37^, sexual and gender minorities^38^, and individuals from low socioeconomic status groups^37^. Importantly, among diverse and underrepresented populations, a HiTOP-consistent (including the overarching *p*-factor), transdiagnostic approach to modeling psychopathology and substance use may be particularly helpful in capturing associations to the pernicious effects of stigma and other minority stressors^39,40^. For example, in a large, U.S. sample of Black Americans and Caribbean-descended Black individuals, cross-sectional reports of perceived racial discrimination were associated with psychiatric disorders as well as Internalizing and Externalizing domains^41^. However, once associations with HiTOP domains were accommodated within the model, few associations with specific disorders remained, suggesting the utility of a transdiagnostic dimensional lens in understanding social determinants of health rather than a disorder-specific approach. Despite the promise of a HiTOP-derived model as applied to diverse and understudied populations, research in this domain is only just beginning to emerge. A critical consideration is the extent to which core domains of the HiTOP model represent universal structures and are measured equivalently across diverse populations and sociodemographic identity groups^40^. There is substantial cross-national research that provides support for the generalizability of specific core domains of the HiTOP model; that is, Internalizing, Externalizing, and Thought Disorder domains^9,40,42,43^. As it pertains to measurement invariance, a handful of studies provide empirical support for the factorial invariance of the model among groups defined by sociodemographic identity (e.g., race and ethnicity^44^, sexual minority identity^45^). The present study assesses the relationship between HiTOP and SUDs in a sample of primarily lower income Black/African American substance users in treatment, as well as in a nationally representative sample (National Comorbidity Study-Replication, NCS-R)^46^. Thus, this study will allow for a replication of the fit of HiTOP models across these populations, as well as assessment of the relationship between SUDs and psychopathology within and between these populations.

### Current Study

The current study aims to develop an understanding of how SUD/PSUD relates to the general psychopathology (*p*) factor and key spectra of the HiTOP model as conventionally constructed (based on the available diagnoses). These key spectra included: INT (including Fear and Distress subfacets), EXT (including SUDs and Antagonism subfacets), and TD (with no subfacets). First, the HiTOP model was applied to both a sample of primarily lower income treatment seeking substance users who are primarily Black/African American, and a nationally representative sample (NCS-R), to verify the fit of HiTOP in both samples. Second, we assessed the fit of a model removing SUDs from HITOP in the two datasets, to verify the fit of the model excluding SUDs relative to other psychopathology. For parsimony in the hierarchy (e.g., having all subfacets at the same level), we implemented a model with the following latent measures: Fear, Distress, Antagonism, and the *p*-factor representing the shared variance of the others. To represent SUDs variance in a single variable, we created a SUDs/PSUDs latent factor from the indexed SUDs diagnoses. Next, we assessed the direct relationship between *p*-factor and SUD/PSUD, as well as the individual direct relationships between the subfacets (TD, Fear, Distress, and Antagonism) and SUD/PSUD. Finally, we assessed indirect relationship of TD, Fear, Distress, and Antagonism (as four separate models) on PSUD/SUD, mediated via *p*-factor. The primary hypothesis is that *p*-factor, and lower-order factors, will be significantly related to SUD/PSUD, and that *p*-factor will account for the relationship between SUD/PSUD and INT, EXT, and TD. Because *p*-factor is understood as a measure of problem severity, indexing functional impairment^42^, significant associations of SUD/PSUD with *p*-factor and SUD/PSUD with lower-order measures of psychopathology (i.e., TD, Fear, Distress, and Antagonism) would provide a detailed statistical characterization of the relationship between SUDs and HiTOP transdiagnostic factors.

## Method

### Treatment Center

#### Participants

2617 participants were recruited from an addiction treatment center in the Washington D.C. area. The center is a 160-bed residential treatment facility which serviced local community with limited economic resources. Clients at this facility reported experiencing meeting criteria for substance dependence to several different substances (alcohol, cannabis, opiates, cocaine, hallucinogens, stimulants, sedatives)had varying lengths of stay (e.g., 30 days, 90 days),and were often court-mandated to complete treatment (i.e., undergoing treatment as part of pre-trial processes or following sentencing). The participants were mostly lower-income (mean yearly income < $10,000), non-White racial minority (97% racial/ethnic minorities), interaction with the criminal justice system (87% previously incarcerated, 66% court mandated treatment), education level (38% below high school level; mean education level: GED), involvement with Child Protective Services, and many were non-primary caregivers.

#### Structured Clinical Interview DSM-IV

Generally, within the first week of entering the facility, a subset of clients completed the semi-structured Structural Clinical Interview based on the DSM-IV (SCID-IV)^47^. The clients were selected either based on recommendation from the staff or, if no recommendations were made, based on admittance date (more recently admitted). Among those who were admitted recently, if there was overlap in the dates, a client was selected at random. The clinical interviews were conducted in a private room designated by the staff within the facility. After collecting clinical consent, questions were asked to gather demographic data, treatment history, previous substance use, and SCID-IV questions (diagnosis questions related to substance use, depression, anxiety, etc.). Research consent was requested after completion of the interview. If they did not provide research consent, the data was only used by the facility for treatment purposes and was not used for research, in accordance with the approved Institutional Review Board protocol.

#### Data Organization

For each diagnosis (e.g., depression, anxiety, etc.), data were re-coded as binary 0 (did not meet) or 1 (did meet). Substance use was coded as binary 0 (did not meet) or 1 (met for either Dependence or Abuse). For substances that were less common (sedatives, hallucinogens/PCP, stimulants), a low base rate binary variable was developed to indicate whether a participant met for Abuse or Dependence for at least one (coded as 1) or none (coded as 0) of these substances. For EXT, which generally includes separable Disinhibition and Antagonism factors, we narrowed this to SUDs and Antagonism. The reason for not having a separable Disinhibition subfacet is as follows. Following HiTOP conventions, conduct disorder and antisocial personality disorder (ASPD) cross-load on Antagonism and Disinhibition factors, while SUDs only loads on Disinhibition, and borderline personality disorder (BPD) is only loaded on Antagonism and not Disinhibition. To most appropriately represent the HiTOP model, we included SUDs to index Disinhibition and the others we labeled as Antagonism. Then, when SUDs was removed, it left only Antagonism loading onto EXT, so we removed the EXT level and operated at the subfacet level of Antagonism to avoid redundancy in the latent factors (i.e., EXT and Antagonism would be represented by the same measured diagnosis variables, otherwise). To have all subfacets operate at the same level, we dropped the INT subfacet, and directly included Fear and Distress subfacets at the same level as Antagonism, and TD.

Thus, the two models needed for the present project were constructed as follows. The first conventional model included INT (including Fear and Distress subfacets), EXT (including SUDs and Antagonism subfacets), TD (with no subfacets), and the *p*-factor. The second model removed SUDs and created five latent factors, as follows: Fear, Distress, Antagonism, and TD, as well as the *p*-factor (indexing the shared variance across the other four). The SUDs/PSUDs latent variable was created using measured variables for alcohol use, opioid use, cannabis use, and low base substances of hallucinogen/PCP, stimulant, and sedative use (collapsing across Abuse and Dependence as noted above). All latent factors were computed in Mplus^48^.

### The National Comorbidity Survey-Replication (NCS-R)

#### Participants

NCS-R is a nationally representative household survey of mental disorders in the United States. The NCS-R included a nationally representative sample of adults aged 18 and older. Interviews were conducted between February 2001 and April 2003^49^. The sample was selected using a multistage stratified cluster sampling design representative of the US household population^49^. Our sample was weighted to be representative of the U.S. population^49^; 72.8% non-Hispanic whites, 53% female). Most participants had 13+ years of education (50.7%) and lived in metropolitan areas (68.2%). The NCS-R had a response rate of 70.9%.

#### Measurements

The NCS-R included questions about the prevalence, severity, and impairment associated with a variety of mental health disorders, including INT (e.g., anxiety disorders and depression), EXT (e.g., SUDs and antisocial personality disorder), and TDs (e.g., schizophrenia and mania)^49^.

The survey was administered in two parts. The first section, which was given to all participants (N=9,282), covered all the main mental health disorders that the survey was designed to assess. The second section was only administered to those who met lifetime diagnostic criteria for any mental health disorders and an additional probability sample (*N*= 5,692). It aimed to assess additional disorders and other correlates (e.g. risk factors) (Kessler et al. 2004). Because some of the disorders included in our analysis were addressed in Part II, only Part II participants were included in our sample.

#### Data Organization

We constructed the same two HiTOP-based models for the NCS-R data as for the treatment center data, although the specific indicators were slightly different due to the data available. All latent models were created in Mplus, as was done with the treatment center data. The NCS-R collected DSM-IV (Diagnostic and Statistical Manual of Mental Disorders-IV)^50^ and ICD-10 (International Classification of Diseases-10)^51^ diagnostic data from participants, in addition to other mental health information^52^. For example, the Fear subfacet included panic disorder, obsessive-compulsive disorder (OCD) and social phobia available in the treatment center data, but added agoraphobia, specific phobias, separation anxiety disorder (SEPAD) available in the NCS-R data. Distress had mostly the same indicators, with the addition of Dysthymia in the NCS-R data. The SUDs latent variable was computed from the shared variance between drug abuse, drug dependence, nicotine dependence, alcohol abuse and alcohol dependence diagnoses available in the DSM-IV and ICD-10 codes.

Notably, certain disorders in our model -- namely, BPD, ASPD, OCD, and psychosis disorders --, were not included in the NCS-R’s diagnostic section. To address this, the variables for these diagnoses were constructed by identifying NCS-R survey questions that corresponded to the SCID-5 interview questions for each disorder. We employed two coding strategies for these variables: a categorical approach and a continuous approach. The categorical approach (used for the psychosis and OCD variables) categorized individuals as having the disorder if they had responded “yes” to any of the questions selected, which were diagnostic in nature. For instance, if an individual responded affirmatively to “Ever hear voices when not dreaming/asleep/under the influence of substances?”, they were considered to have a psychosis disorder. The continuous approach (used for ASPD and BPD) assigned a score based on the number of “yes” responses, so that a higher number of affirmative responses meant a higher score, indicating a higher presence of the disorder. This approach was necessary for personality disorders because the NCS-R questions were behavioral in nature, therefore, a single affirmative response to a personality disorder question was insufficient for a diagnosis. A continuous approach was used based on a cumulative score for personality disorders, since the diagnosis of a personality disorder necessitates a constellation of behaviors or symptoms, it cannot be captured by endorsing a single item. All indicator data was organized, and variables created for analysis, using RStudio^53^.

### Statistical Analyses

For both datasets, structural equation modeling was conducted using Mplus 8.7^48^. The analyses included confirmatory factor analyses to identify latent factors of the HiTOP model and polysubstance use (represented as the SUDs latent variable measured by multiple SUDs), and the regression pathways to relate these latent factors. The model tested both direct and indirect relations (i.e., mediated pathways) using a weighted least squares mean and variance adjusted estimator. This estimator was selected because it is robust against non-normal data and suitable for handling the categorical variables included in the model. The model fit will be assessed by the following Mplus consistent indices (with mentioned cutoffs): Comparative Fit Index (CFI; good fit ≥.95, acceptable fit ≥.90), Tucker-Lewis Index (TLI; good fit ≥.95, acceptable fit ≥.90), Standardized Root Mean Square Residual (SRMR; good fit ≤.08), Root Mean Square Error of Approximation (RMSEA; good fit ≤.06, acceptable fit ≤.08)^54^. Chi-square values are also included; however given the large sample size, greater emphasis is placed on the aforementioned fit measures^55^.

Hypotheses for the conventional HiTOP model was that the model would fit both datasets. For the second model, assessing SUDs, the hypothesis was first that the models would similarly fit both datasets. Second, we hypothesized that SUDs would be related to *p*-factor and each of the subfacets (direct effects), and that some unique SUDs variance would be related to the subfacets beyond the *p*-factor (indirect effects).

**Figure 1.**
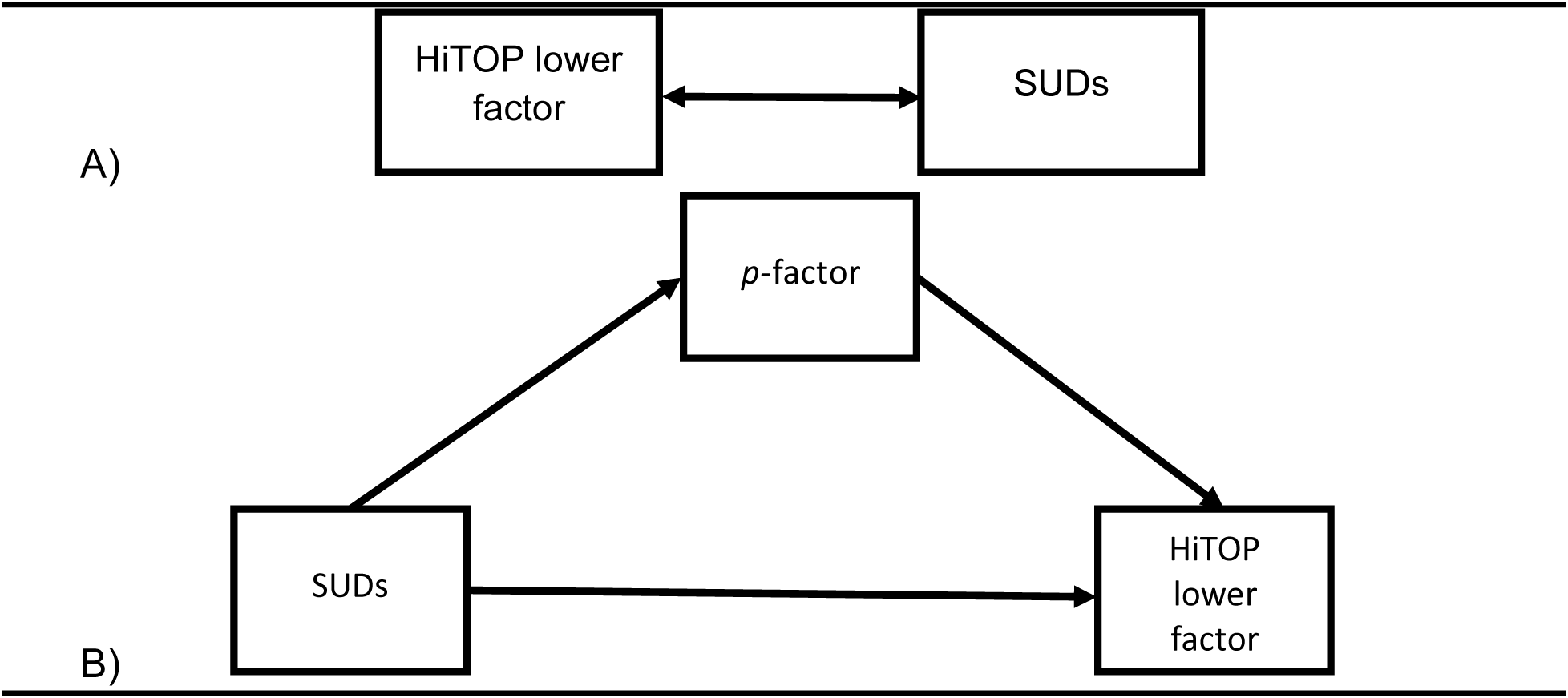
Conceptual visualization of the direct (A) and indirect (B) effect models. HiTOP lower order factor refers to TD, Fear, Distress, and Antagonism, and each of the four factors were reviewed for the direct relationship with SUDs and their indirect relationship with SUDs after mediating for *p*-factor.

## Results

### Replicating HiTOP

The first step was to replicate the conventional HiTOP model in both datasets.

#### Treatment Center

The information is displayed below in Figure 2 and presented with reasonable fit: ***χ*^2^ =425.287 (*df*=96,*p*<.001), CFI = .931, TLI = .914, SRMR = .067, RMSEA = .036 (.033, .040)**. The model presents with good fit for SRMR and RMSEA, and acceptable fit for CFI and TLI.

**Figure 2.**
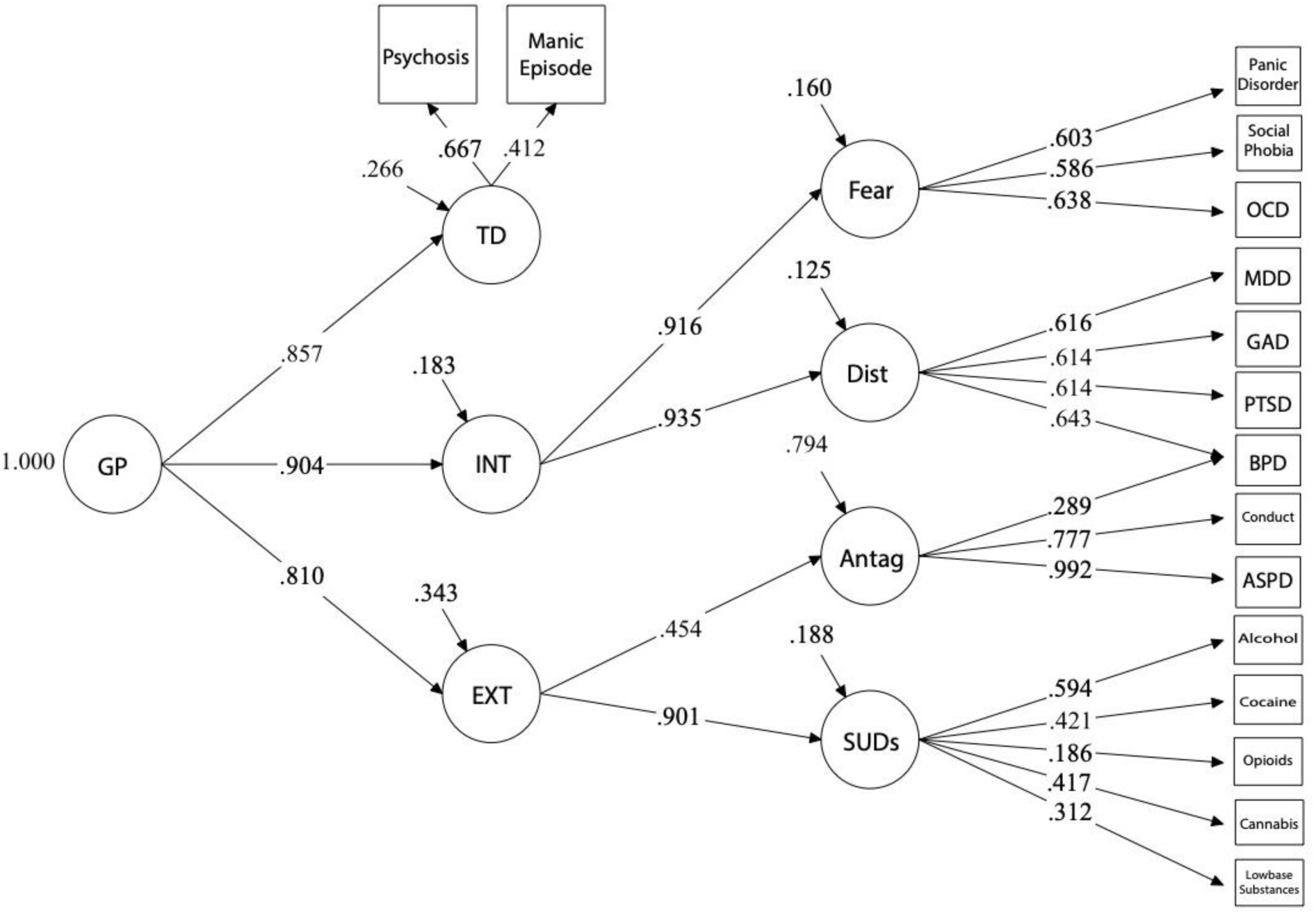
SCID-IV Conventional HiTOP model. *Note.* Abbreviations for all models and tables: GP is General Psychopathology factor, TD is Thought Disorder, INT is Internalizing, EXT is Externalizing, Dist is Distress, Antag is Antagonism, SUDs is Substance Use Disorders, OCD is Obsessive Compulsive Disorder, MDD is Major Depressive Disorder, GAD is General Anxiety Disorder, PTSD is Posttraumatic Stress Disorder, BPD is Borderline Personality Disorder, and ASPD is Antisocial Personality Disorder.

#### NCS-R

The information is displayed in Figure 2.1 and presented with reasonable fit: ***χ*^2^ = 1083.903 (df=222,p<.001), CFI = .970, TLI = .966, SRMR = .056, RMSEA = .026 (.025, .028).** The model presents with good fit for CFI, TLI, SRMR, and RMSEA.

**Figure 2.1.**
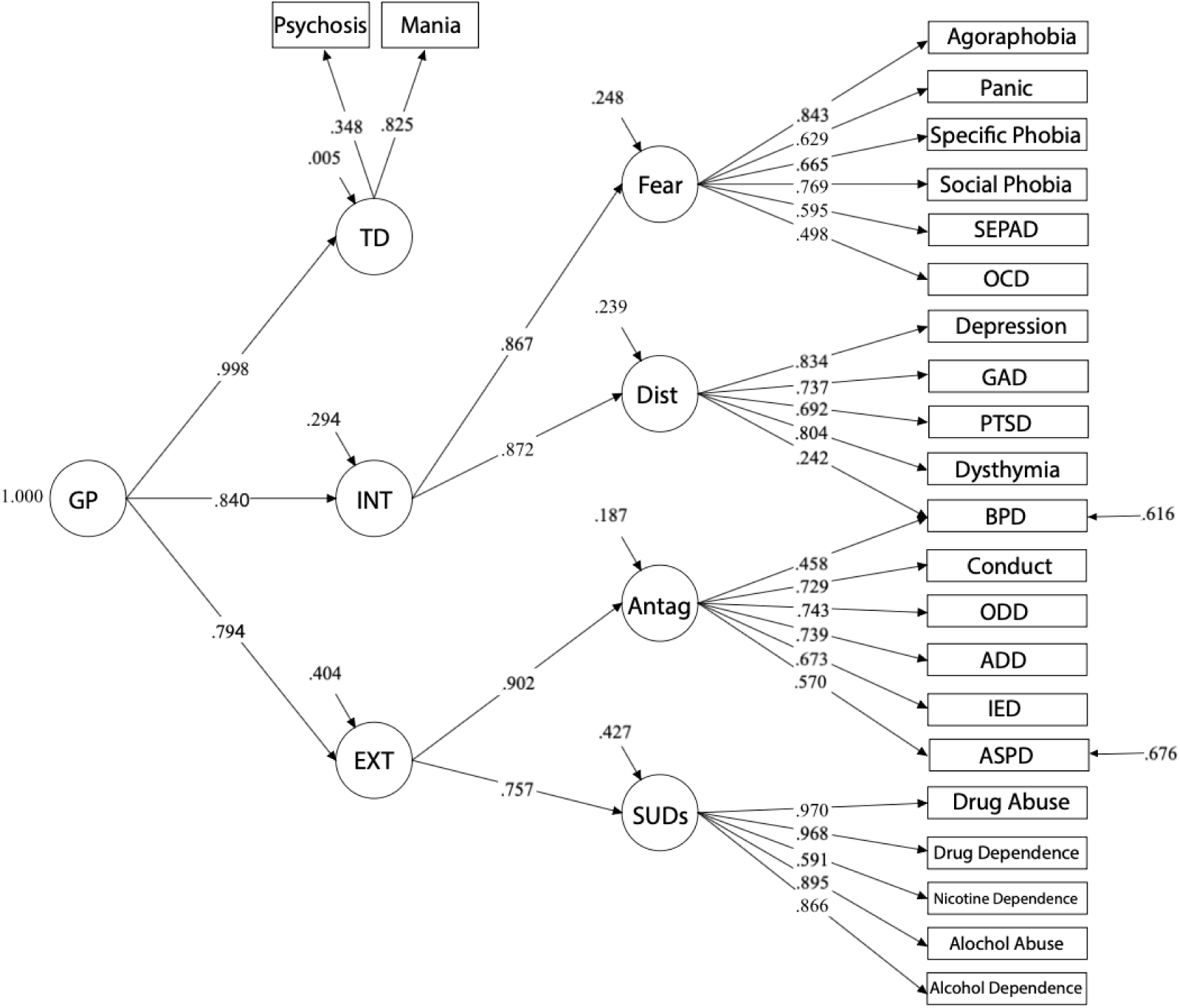
NCS-R Conventional HiTOP model. Residual variance is observed for ASPD and BPD because these constructs were modeled as continuous variables. Abbreviations not addressed in Figure 2 note: SEPAD is Separation Anxiety Disorder, ODD is Oppositional Defiant Disorder, ADD is Attention Deficit Disorder, and IED is Intermittent Explosive Disorder.

### HiTOP Model without SUDs

Next, given SUDs relationship with both INT and EXT disorders, it was important to remove SUDs from the development of *p*-factor to interpret its direct and indirect relationships with *p*-factor and its lower-order factors (i.e., TD, Fear, Distress, Antagonism). Additionally, the INT and EXT levels were removed because after removing SUDS, EXT would only be built of Antagonism in this model, which is effectively the same as looking at the direct loading of Antagonism onto *p*-factor. This lower level also allows us to look at Fear and Distress, separately.

#### Treatment Center

After removing SUDs, the current HiTOP model presented with these fit indices: ***χ*^2^ =107.784 (*df*=39, *p*<.001), CFI = .981, TLI = .974, SRMR = .063, RMSEA = .026 (.020, .032)**. The model presents with good fit for CFI, TLI, SRMR, and RMSEA.

**Figure 3.**
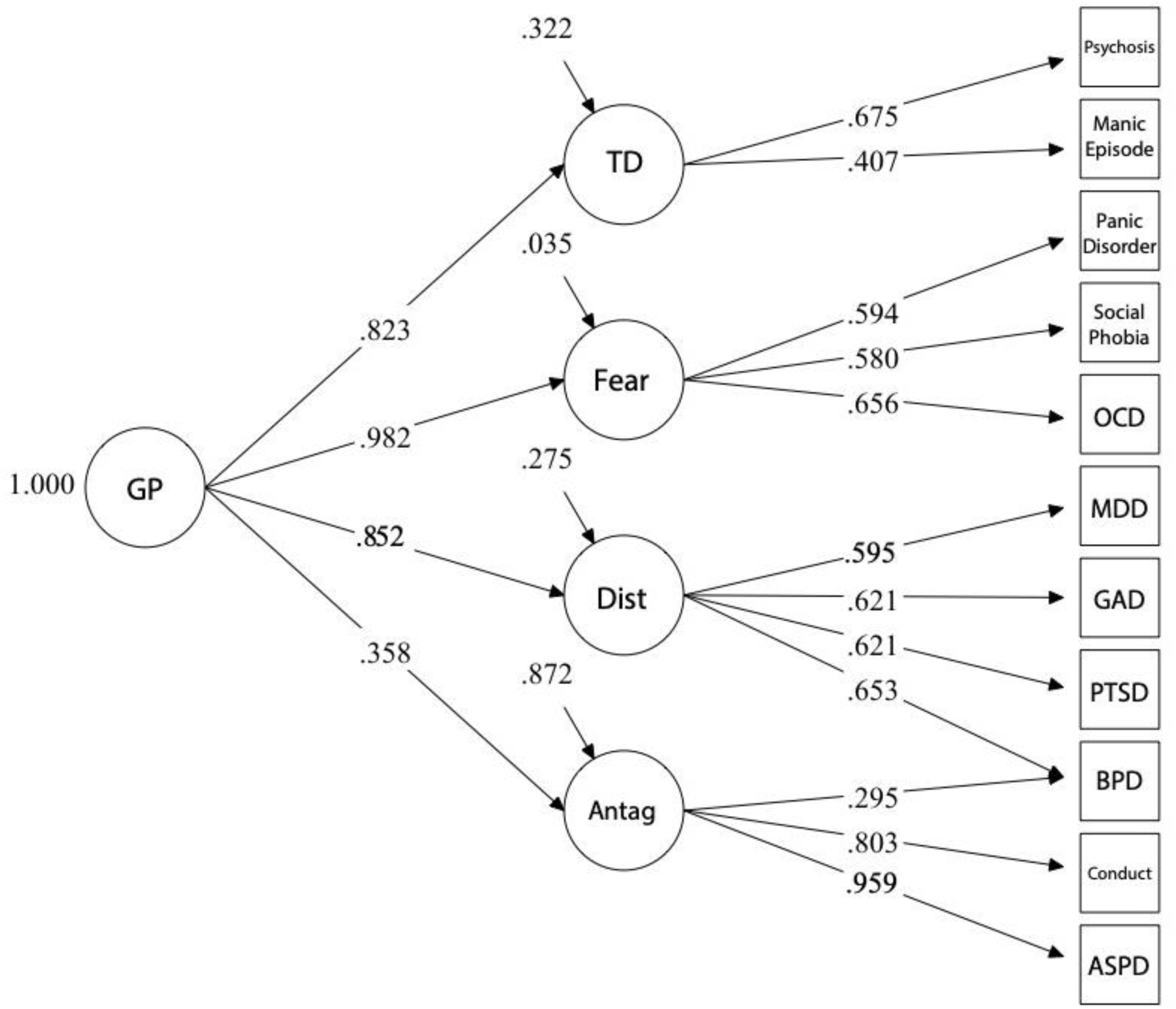
SCID-IV HiTOP model removing SUDs factor.

#### NCS-R

After removing SUDs, the current HiTOP model presented with these fit indices: ***χ*^2^ = 851.150 (df=130, p<.001), CFI = .956, TLI = .948, SRMR = .058, RMSEA = .031 (.029, .033).** The model presents with good fit for CFI, TLI, SRMR, and RMSEA.

**Figure 3.1.**
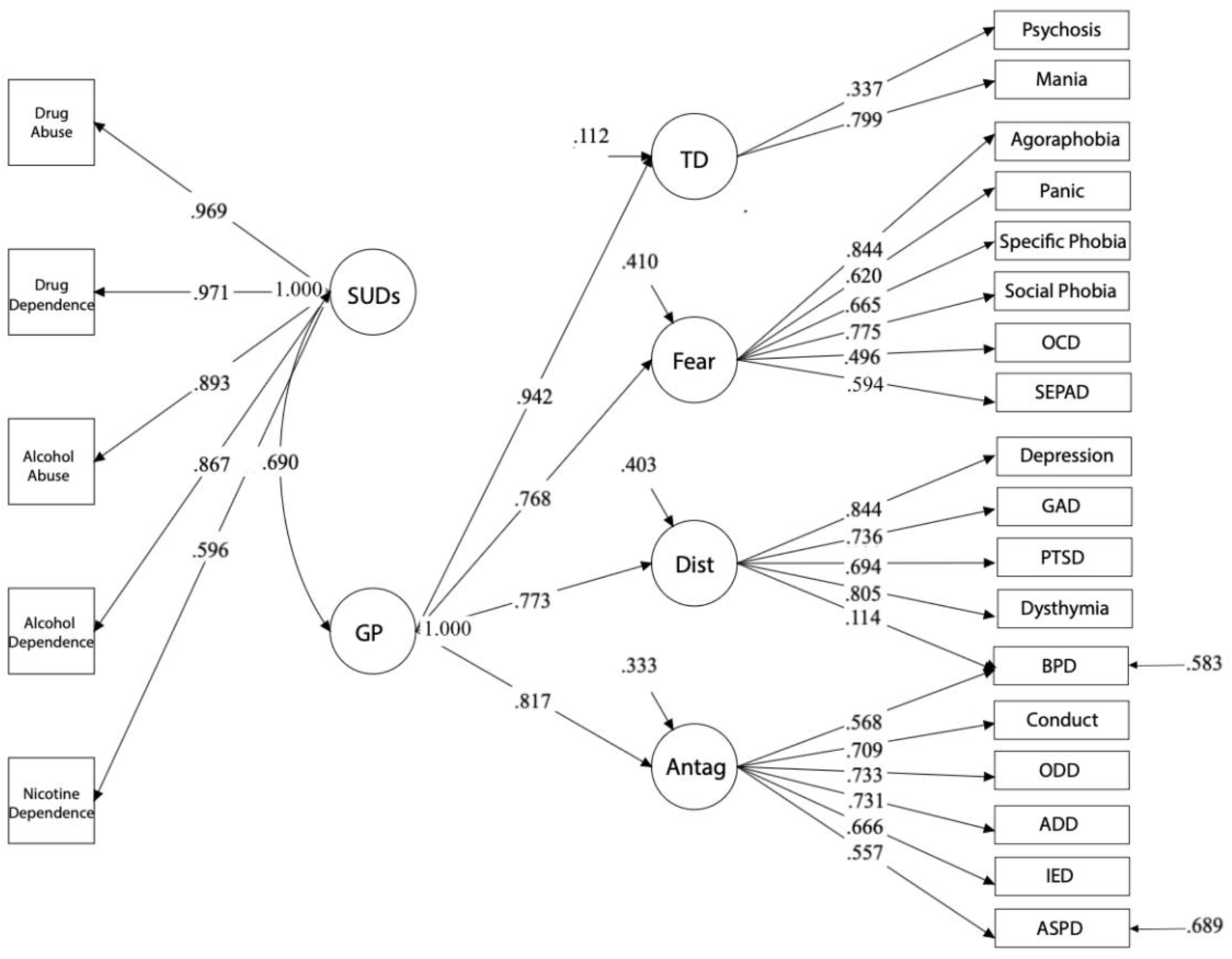
NCS-R HiTOP model removing SUDs factor. Residual variance is observed for ASPD and BPD because these constructs were modeled as continuous variables.

### Individual SUDs Measures and p-factor

#### Treatment Center

Next, individual substance use measures were related to *p*-factor through a point-biserial correlation to better understand the impact of individual substance usage compared to comorbid substance usage. The following variables were developed from existing substance use data: Alcohol Only Ever, Cannabis Only Ever, Cocaine Only Ever, Opioid Only Ever, and Low-base Only Ever. Each of these variables was a binary measure with 0 being not meeting for any substance at all, 1 being meeting only for the respective substance but no other substances, and all other values being considered missing data. Below is a summary of the results from this analysis.

**Table 1.** Treatment Center: Individual SUDs Correlated with *p*-factor.

|  | *N*=2617 | *r* |
| --- | --- | --- |
| Alcohol Only Lifetime | 0=384<br>1=147<br>Missing=2086 | .481*** |
| Cannabis Only Lifetime | 0=384<br>1=89<br>Missing=2144 | .281* |
| Cocaine Only Lifetime | 0=384<br>1=308<br>Missing=1925 | .551*** |
| Opioid Only Lifetime | 0=384<br>1=141<br>Missing=2092 | .352*** |
| Low-base Only Lifetime | 0=384<br>1=304<br>Missing=1929 | .739*** |

† .10≤*p*<.05, \**p*≤0.05, \*\**p*≤0.01, \*\*\**p*≤0.001.

#### NCS-R

For the NCS-R data, three variables were developed: Combined Drug Only Ever, Alcohol Only Ever, and Nicotine Only Ever. For Combined Drug Only Ever and Alcohol Only Ever, the variables were coded as 1 if either Abuse or Dependence was met. All variables were coded as 0 if the participant met for no substances. Remaining participants were coded as missing for the variable.

**Table 1.1.** NCS-R: Individual SUDs Correlated with *p*-factor.

|  | *N*=5692 | *r* |
| --- | --- | --- |
| Combined Drug Only Lifetime | 0=4177<br>1=93<br>Missing=1422 | .407*** |
| Alcohol Only Lifetime | 0=4177<br>1=402<br>Missing= 1113 | .396*** |
| Nicotine Only Lifetime | 0=4517<br>1=359<br>Missing=816 | .382*** |

† .10≤*p*<.05, \**p*≤0.05, \*\**p*≤0.01, \*\*\**p*≤0.001.

### HiTOP Related to SUDs

Next, the relationship between SUDs and *p*-factor is demonstrated. A correlational relationship was measured to simply identify that a relationship between the two measures existed. Given this relationship (presented below), the direct and indirect relationships between lower-order factors and SUDs can be analyzed.

#### Treatment Center

There is a strong relationship between SUDs and *p*-factor at .761. This model relating p-factor to SUDs presents with reasonable fit: ***χ*^2^ =444.898 (*df*=98, *p*<.001), CFI = .927, TLI = .911, SRMR = .069, RMSEA = .037 (.033, .040)**. The model presents with good fit for SRMR and RMSEA, and acceptable fit for CFI and TLI.

#### NCS-R

SUDs and *p*-factor demonstrate a strong relationship at .690. This model relating *p*-factor to SUDs presents with a reasonable fit: ***χ*^2^ =1678.386 (df=224, p<.001), CFI = .950, TLI = .943, SRMR = .071, RMSEA = .034 (.032, .035).** The model presents with good fit for CFI, SRMR, and RMSEA, and acceptable fit for TLI.

### Direct and Indirect Effects of SUDs on HiTOP

The direct and indirect relationships were analyzed with a constrained HiTOP model. The constrained model restricted the loadings to match all of the loadings identified in the model in Figure 4 and Figure 4.1, which holds the interpretation of the latent variables constant between the two samples.

**Figure 4.**
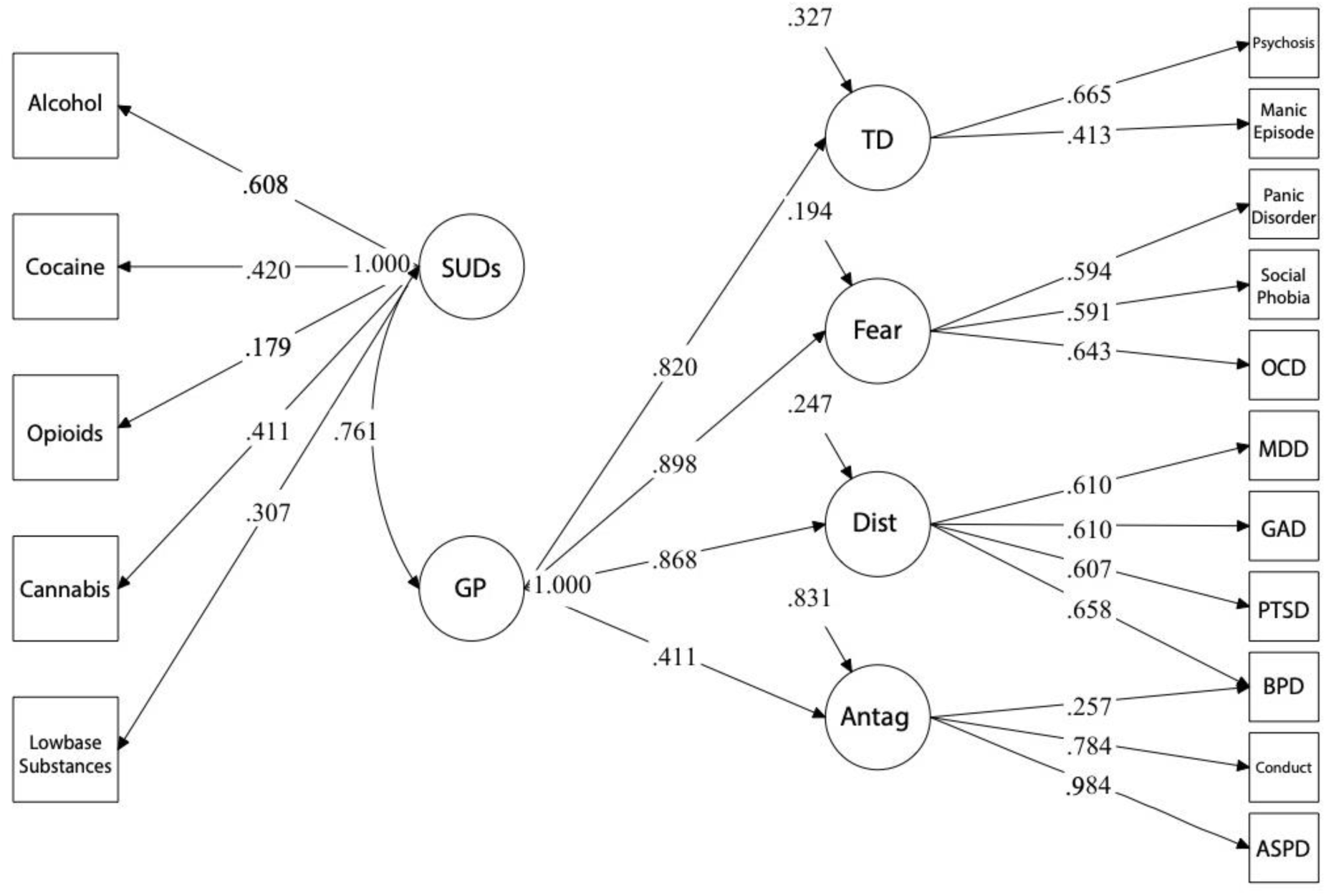
SCID-IV *p*-factor covarying with SUDs.

**Figure 4.1.**
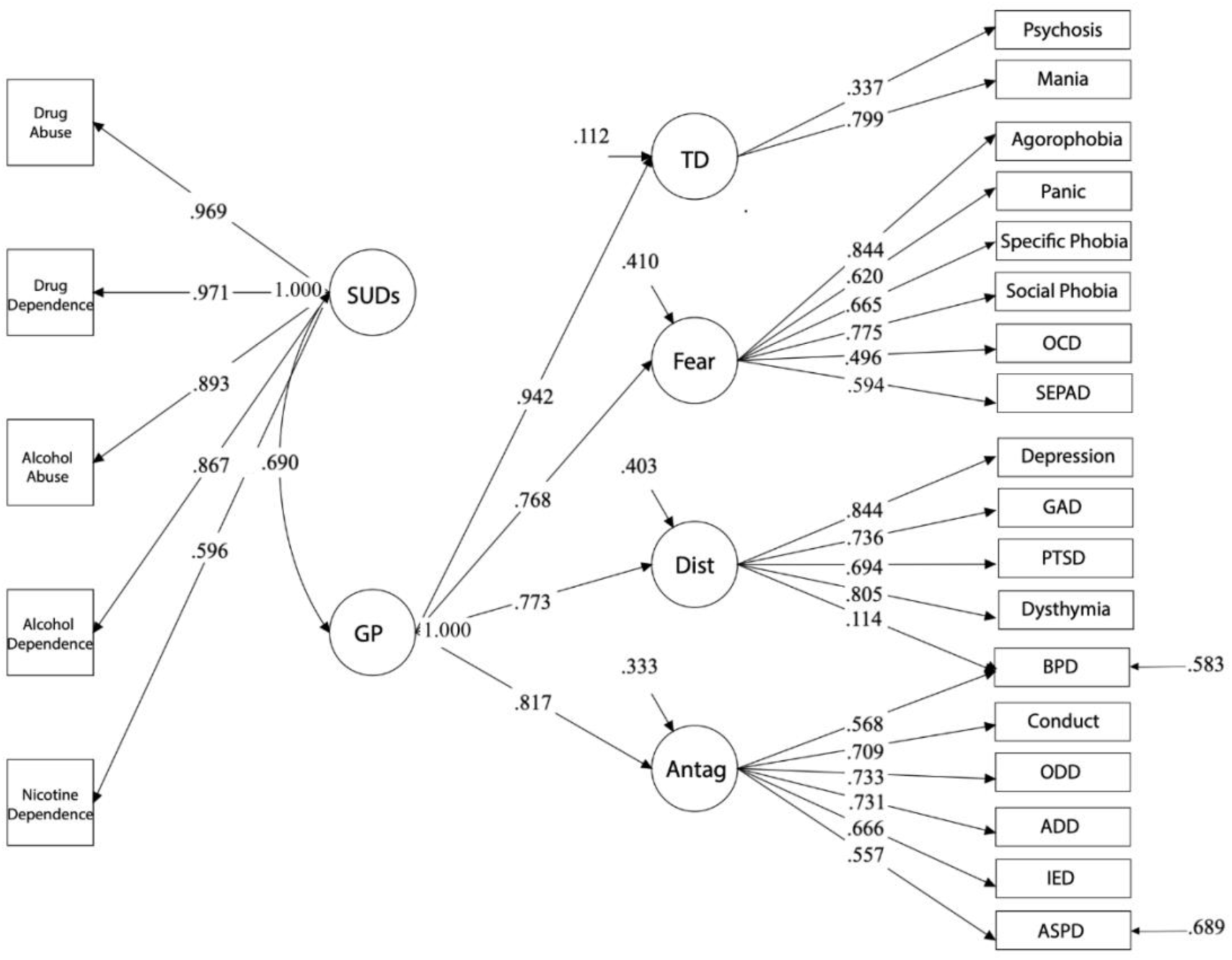
NCS-R *p*-factor covarying with SUDs. Residual variance is observed for ASPD and BPD because these constructs were modeled as continuous variables.

#### Treatment Center

In the constrained model, there were strong direct effects between SUDs and all the lower order measures of HiTOP. However, when looking at the indirect effect between SUDs and the lower order measures (i.e., *p*-factor mediating the relationship between SUDs and respective lower order measure), the relationship between SUDs and TD and SUDs and Distress were no longer significant. Although the relationship between Antagonism and SUDs was not as strong, there was still a moderate relationship between the two measures after accounting for *p*-factor (.318). Notably, the relationship between SUDs and Fear became a strong negative relationship after accounting for the variance from *p*-factor (−.648).

**Table 2.1.** Treatment Center: Constrained *p*-factor loadings related to lower-order HiTOP measures.

| SUDs Regressed on... | Direct | Indirect |
| --- | --- | --- |
| TD | .760*** | -.055 |
| Antag | .659*** | .318*** |
| Fear | .758*** | -.648*** |
| Dist | .734*** | .025 |

† .10 ≤ *p* < .05, \**p* ≤ 0.05, \*\**p* ≤ 0.01, \*\*\**p* ≤ 0.001.

#### NCS-R

All direct effects were statistically significant and positive. In comparison, the indirect effects were overall weaker and the relationships between SUDs and TD, SUDs and Fear, and SUDs and Distress were no longer significant. Notably, while TD, Fear, and Distress, initially demonstrated positive direct effects (.855, .669, .653, respectively), TD and Fear exhibited negative, nonsignificant indirect effects (−.041, −.067, respectively) and Distress continued to exhibit a positive, though nonsignificant, indirect effect (.012). Additionally, there remained a significant positive relationship between Antagonism and polysubstance use (.354).

**Table 2.2.** NCS-R: Constrained *p*-factor loadings related to lower-order HiTOP measures.

| SUDs Regressed on... | Direct | Indirect |
| --- | --- | --- |
| TD | .855*** | -.041 |
| Antag | .699*** | .354*** |
| Fear | .669** | -.067 |
| Dist | .653*** | .012 |

† .10 ≤ *p* < .05, \**p* ≤ 0.05, \*\**p* ≤ 0.01, \*\*\**p* ≤ 0.001.

## Discussion

The present study provides new information about the relationship between SUDs and the HiTOP transdiagnostic framework. A pivotal aspect of our study involved the removal of SUDs from the EXT spectrum in our HiTOP model, allowing modeling of the relationship to all core dimensions of the HiTOP model. This includes the overarching *p*-factor and the relationship for the lower order dimensions (INT, EXT, and TD). The primary results from this study indicate that there is a robust positive relationship between SUDs and the overarching *p*-factor, providing a parsimonious representation of a majority of variance in each latent factor. Significant indirect effects indicate differential independent contributions of the lower order dimensions. By creating a SUDs latent factor, the findings have relevance across substances, and support the view that an increasing number of substances used (polysubstance use) is related to increased psychopathology. Finally, the primary findings replicated conceptually across a nationally representative sample and a sample representing primarily patients with lower SES in inpatient treatment who identify as racial/ethnic minorities (primarily African American). Together findings from this study support several inferences. First, SUDs are broadly associated with psychopathology, supporting the view that treating psychopathology may be central to treating SUDs. Second, in addition to an overall positive relationship, INT and EXT may also contain some inhibitory and facilitatory mechanisms respectively, while TD showed no additional prediction of SUDs beyond the *p*-factor. Finally, these findings provide additional support for the development of interventions aimed at the level of transdiagnostic dimensions.

### Limitations of the *p*-factor

A recent critique has brought needed scrutiny to the idea of a broad overarching latent transdiagnostic psychopathology measure, i.e., the *p*-factor^56^. The criticisms include: poor replication of *p*-factor measures, poor validation, and lack of support for the *p*-factor as a latent causal entity. These criticisms are important and draw into question what inferences are appropriate relative to the *p*-factor. What fits within these critiques is the idea that the *p*-factor represents an index of functional impairment, because DSM diagnoses fundamentally integrate functional impairment into the criteria. Thus, we limit our inferences about the *p*-factor from the current findings to that of functional impairment or psychopathology severity, consistent with limitations suggested by Watts^56^.

In the context of these issues, we also note the conceptual replication of the present effects across two importantly different datasets (nationally representative and primarily African American substance users in treatment), with different variables making up the model of the *p*-factor. This suggests that the observed effects are robust across different *p*-factor measures from different populations, bolstering confidence that the findings are reliable and generalizable.

### Implications for the Relationship Between SUDs/polySUDs and *p*-factor

Across local and national samples the SUDs/PSUDs measures were robustly positively associated with the *p*-factor. Broadly, this relationship has long suggested that substance use is associated with increased functional impairment central to psychopathology^49,57,58^.

PolySUDs is broadly associated with a variety of psychopathology domains, spanning Thought Disorder, Antagonism, Fear, and Distress—with similar associations across the board. This is consistent with this relationship potentially being accounted for by a general factor of psychopathology.

As previous work indicates and as is shown in this study, there exists a co-occurrence of mental health and SUDs that is well measured^6^; however, treatment is not as well geared towards this co-occurrence as may be beneficial. In a survey of 7.7 million adults with cooccurring mental health and SUDs, only 9.1% received treatment for both^59^. Additionally, 34.5% received only mental health care and 3.9% received only substance use treatment, with the remaining 52.5% receiving care for neither. A primary barrier is that only about 15% of centers specialize in treating both at their center^60^. This finding provides evidence that, given this strong relationship, it is compelling to address them concurrently.

Additionally, the relationship of this SUDs/PSUDs measure with *p*-factor aligns with the notion that polysubstance use, compared to individual SUDs, will be related to increases in the *p*-factor. Polysubstance use has been shown to have worse outcomes than individual SUD diagnoses^61^. The correlations here depict that one reason for this may be due to a stronger relationship to psychopathology for comorbid use when compared to individual use for any of the substances measured here (i.e., alcohol, cannabis, cocaine, opioids, or low-base rate substances of sedatives, stimulants, and hallucinogens). This can mean that it would be beneficial to tailor treatments to address the underlying transdiagnostic factor of substance use rather than individual substance use.

### Relationship to Lower Order Factors

When controlling for the variance accounted for by the *p*-factor in the current work, some significant residual relationships are observed, suggesting unique contributions, and some nonsignificant residual relationships are observed, suggesting *p*-factor accounts for the SUDs-HiTOP dimension relationship. For TD, the relationship is accounted for by *p*-factor both in the treatment center data and in the national data, indicating that *p*-factor accounts for the variance in TD across samples. For Antagonism (a key subfacet of Externalizing), there remain unique positive associations across both datasets, suggesting a synergistic relationship between SUDs and Antagonism beyond the effect of all other diagnoses. Regarding Internalizing, there are unique indirect negative associations with Fear-related disorders in the treatment center data, only. These differences in Fear-related disorders may be due to variations in data collection methods (e.g., NCS-R may have underdiagnosed or subclinical reporting) and/or differences in sample characteristics across the two datasets (discussed further in the section below). These differences should be evaluated in replication and extension studies. Broadly, indirect effects provide the most support for the idea of Antagonism as an additional risk factor beyond *p*-factor, some support for the Internalizing Fear factor as a protective factor, and suggestive support for TD as a potential small additional risk factor.

### Consistent results across disparate samples

Across two socio-demographically disparate samples (i.e., individuals from a health disparity population compared to a nationally representative sample), the present study provided a conceptual replication of HiTOP model fit with inferences robust across variance in models and model indicators. Further, across both samples, we demonstrated that while HiTOP INT, EXT, and TD dimensions were all related to SUDs, these relationships were partially mediated by their shared variance captured by the *p*-factor, where SUDs and *p*-factor were robustly associated. Notably, in the treatment sample only, unique indirect associations with Fear-related disorders emerged. This is not entirely surprising in light of evidence pointing to common overlapping neural circuits of Fear and problematic drug use (relapse in particular)^62^. More generally, this overlapping neural circuitry may partially account for the high comorbidity of Fear-related and substance use disorders^62^. Overall, the treatment center sample may reflect a population who presents with greater symptom severity, comorbidity, and more complex psychiatric diagnoses. As such, the relevance of *p*-factor as an indicator of functional impairment may be greater in health disparity, treatment-engaged populations.

Among diverse and underrepresented populations, this transdiagnostic approach (as opposed to disorder-specific) to modeling psychopathology and substance use may be particularly useful in identifying pathways from stigma and other minority stressors to mental health outcomes^39–41^. For example, in a national household sample of African Americans and Black individuals of Caribbean descent, Rodriguez-Seijas and colleagues found that transdiagnostic factors—specifically INT and EXT—accounted for the majority of the association between racial discrimination and specific disorders^41^. Regarding substance use in particular, racial discrimination was indirectly related to alcohol and drug use disorders via INT and EXT transdiagnostic factors^41^. Extensions of this work are needed to evaluate the HiTOP and *p*-factor model as a more parsimonious representation of psychopathology with etiological pathways to SUDs across minoritized and majority communities. HiTOP-framed *p*-factor may interact with negative, positive, and regulatory processing and ultimately increase substance use as captured by SUDs. Finally, the discrepancies in measurement and models across the two samples in the current study prevents a formal test of measurement invariance. Future research might build from emerging work assessing the extent to which core domains of the HiTOP model represent universal structures and are measured equivalently across diverse populations and sociodemographic identity groups^40^.

### Limitations and Future Directions

Although these findings provide support for a relationship between polySUDs and *p*-factor, as well as unique relationships with lower order HiTOP measures, there are important limitations that come from this analysis. First, the conceptual replication across different data collections substantiates that the inferences are robust; however, this prevents a formal test of measurement invariance^63^. Without a formal test, the generalizability of the findings should be further explored in other contexts and datasets. Another limitation to consider is the loading of Antagonism onto *p*-factor in the treatment center data. Although the loading meets typically accepted criteria of .3 or higher, the residual variance indicates that a meaningful amount of the variance is not accounted for in the model. Further analysis in similar treatment center settings would be beneficial to better understand this limitation. Despite this measurement difference between the two datasets in the current study (i.e., NCS-R data does not have the same concern), we do still see similar results across the two models.

From this work, there are a few next steps that would be beneficial to pursue. First, looking at these association longitudinally will allow for more predictive analyses than could be made from the current correlational study. This longitudinal work could also focus on potential risk factors and outcomes related to SUDs and psychopathology^8,64^. Additionally, considering the commonalities between antagonistic and SUDs behaviors^65^, future work should consider separating the two more often to understand unique downstream consequences like role and relationship impairment. This separation can better inform conceptual understandings, as well as practice in developing more appropriate therapeutic methods.

### Conclusions

The present findings add to evidence that substance use and mental health are fundamentally related, and that effectively addressing substance use, will involve addressing mental health concerns. This interconnection was observed in both minoritized and representative samples, suggesting this relationship is pervasive in the population. Finally, these findings provide additional motivation for developing transdiagnostic treatment targets.

## Supporting information

Supplemental Unconstrained Model Results

## Data Availability

NCS-R data reviewed in the present study is publicly available online at https://www.icpsr.umich.edu/web/ICPSR/studies/20240.
Remaining data reviewed in the present study is available upon reasonable request to the authors.

