## Supplemental Unconstrained Model Results for "General psychopathology and substance use disorders: An indicator of functional impairment"

### Supplemental Data

#### Unconstrained Models

**Table S1.** Treatment Center: Unconstrained *p*-factor loadings related to lower-order HiTOP measures.

| SUDs Regressed on… | Direct | Indirect |
| --- | --- | --- |
| TD | .763*** | -.054 |
| Antag | .659*** | .318*** |
| Fear | .758*** | -.649*** |
| Dist | .734*** | .022 |

**Table S1.1.** NCS-R: Unconstrained *p*-factor loadings related to lower-order HiTOP measures.

| SUDs Regressed on… | Direct | Indirect |
| --- | --- | --- |
| TD | .488*** | -.196*** |
| Antag | .534*** | .485*** |
| Fear | .463*** | -.564*** |
| Dist | .514*** | -.367*** |
